# The contribution of genetic risk to the comorbidity of depression and anxiety: a multi-site electronic health records study

**DOI:** 10.1101/2022.04.11.22273720

**Authors:** Brandon J Coombes, Isotta Landi, Karmel W Choi, Kritika Singh, Y Nina Gao, Brian Fennessy, Greg D Jenkins, Anthony Batzler, Richard Pendegraft, Nicolas A Nunez, Euijung Ryu, Priya Wickramaratne, Jyotishman Pathak, J John Mann, Lea K Davis, Jordan W Smoller, Mark Olfson, Alexander W Charney, Joanna M Biernacka

## Abstract

**Importance:** Depression and anxiety are common and highly comorbid, posing a clinical and public health concern because such comorbidity is associated with poorer outcomes.

**Objective:** To evaluate association of genetic risk scores with depression and anxiety diagnosis either in isolation or comorbid with each other.

**Design:** International Classification of Diseases (ICD) ninth and tenth edition codes were extracted from longitudinal electronic health records (EHR) from four EHR-linked biobanks with genetic data available. Data analysis was performed between February 2021 to October 2021.

**Setting:** EHR-linked biorepository study.

**Participants:** Across the four biobanks, 140947 patients (80601 female [57.2%] including 109592 European ancestry [77.8%], 22321 African ancestry [15.8%], and 9034 Hispanic [6.4%]) were included in the study.

**Main outcomes and measures:** Polygenic risk scores (PRS) for depression and anxiety were computed for all participants. They were assessed for diagnosis of depression and anxiety using ICD9/10 codes. The primary outcome was a four-level depression/anxiety diagnosis group variable: neither, depression-only, anxiety-only, and comorbid. Estimated effect measures include odds ratios and the proportion of variance on the liability scale explained by the PRS.

**Results:** 95992 patients had neither diagnosis (68.1%), 14918 depression-only (10.6%), 12682 anxiety-only (9.0%), and 17355 comorbid (12.3%). PRS for depression and anxiety predicted their respective diagnoses within each biobank and each ancestry with the exception of anxiety-PRS not predicting anxiety in any ancestral group from one biobank. In the meta-analysis across participants of European ancestries, both PRSs for depression and anxiety were higher in each diagnosis group compared to controls. Notably, depression-PRS (OR=1.20 per SD increase in PRS; 95% CI= 1.18-1.23) and anxiety-PRS (OR=1.07; 95% CI=1.05-1.09) had the largest effect size for the comorbid group when compared to controls. The confidence interval for the depression-PRS effect did not overlap across groups demonstrating a gradient of genetic risk across the four diagnosis groups.

**Conclusions and Relevance:** The genetic risk of depression and anxiety make distinct contributions to the risk of comorbid depression and anxiety, supporting the hypothesis that the correlated disorders represent distinct nosological entities.

**Key Points:** *Question:* Is the genetic risk of depression and anxiety associated with comorbidity of depression and anxiety?

*Findings:* Using electronic health records from four academic medical centers, this study found that genetic risk of depression and anxiety are jointly associated with clinical depression and anxiety diagnoses with better prediction occurring for a diagnosis of depression.

*Meaning:* The genetic risk of depression and anxiety make distinct contributions to comorbid depression and anxiety, which supports the hypothesis that the correlated disorders represent distinct nosological entities.

## Introduction

Depression and anxiety disorders are among the most common psychiatric disorders with a lifetime prevalence of 17% and 30%, respectively.^1^ Symptoms of depression and anxiety often co-occur with nearly half of adults with anxiety also reporting depressive symptoms.^2^ Comorbid depression and anxiety represent a significant clinical and public health concern and is strongly associated with greater severity and earlier age of onset, more severe depression, increased suicidal ideation, poorer antidepressant response, psychosocial impairment, worse course of illness, and increased comorbidity of substance use disorders.^3–6^

The etiologies of comorbid depression and anxiety disorders remain poorly understood. Epidemiological data suggest that depression and anxiety commonly co-occur because they share underlying liabilities or transdiagnostic factors.^7–9^ Latent variable techniques applied to the intercorrelational epidemiological structure of psychiatric comorbidities have consistently identified a broad internalizing factor that includes depression and anxiety disorders.^7–9^ Clinically, depression and anxiety also have significant overlap with anxiety symptoms being incorporated into some diagnostic criteria for depression.^10^ Further, dysregulation of the hypothalamic-pituitary-adrenal axis and the role of common neurotransmitters have been implicated in both.^11,12^ These observations, together with shared clinical responses to similar pharmacological^13^ and psychosocial interventions^14^ as well as shared genetic variation across internalizing disorders, supports the common liability hypothesis of depression and anxiety disorders.^15^

An alternative assumption, the independent liabilities hypothesis, posits that while mood and anxiety have some overlapping risk factors and clinical characteristics, they also have distinct risk factors and characteristics. Using Research Domain Criteria (RDoC), for example, depression is described as a disorder of impaired reward response, learning, and valuation.^16,17^ In contrast, anxiety is described as a dysfunction of threat detection.^16,17^ Risk factors for each disorder appear to be at least partly distinct, arguing against the idea of a fully shared underlying neurobiological liability.^2^ A recent twin study sought to explain both the comorbidity and distinctive nature of the disorders and found that both depression and anxiety have a positive genetic correlation with behavioral inhibition (response to negative stimuli), but only depression has a negative genetic correlation with behavioral activation (response to positive stimuli).^18^ Proposed risk factors associated with the comorbidity of depression and anxiety include female sex, younger age, higher educational level, and childhood trauma^19^ as well as genetics^20^.

Both depression and anxiety are moderately heritable disorders with estimates of 30-40%^21^ and 20-60%^22^, respectively. Genome-wide association studies (GWAS) of depression and anxiety have largely been performed separately and do not explicitly account for the comorbidity.^21–25^ All of these studies find a robust genetic correlation between depression and anxiety with estimates ranging from 80-95%.^26^ However, none of these studies investigated the differences between the two disorders. While GWAS of psychiatric traits have mostly been limited to individuals of European ancestries,^27^ the most recent GWAS of depression and anxiety have added diversity to the samples with the incorporation of East Asian population-based biobanks^24^ and inclusion of African ancestries from the Million Veterans Program, which largely uses electronic health records (EHR) to identify cases and controls.^21,25^ While studies using the EHR may not fully capture patients’ diagnoses as well as those ascertained directly using diagnostic interviews, EHR-linked biobanks allow access to large collections of patients presenting for clinical care. Thus, EHR studies are well-positioned to study real-world diagnoses, including comorbidity between diagnoses.

In this multi-ancestry study, we leveraged four EHR-linked biobanks from the United States to assess the contribution of genetic risk to the comorbidity of depression and anxiety. We first assessed whether polygenic risk scores (PRSs) for depression and anxiety, each derived using results from large GWAS of patients of European ancestries primarily assessed in clinical research studies, were predictive of the respective disorders defined using EHR-based ICD codes. Next, we evaluated how the genetic risk factors for depression and anxiety jointly contribute to the comorbidity of depression and anxiety. If anxiety and depression PRS contribute independently to risk of comorbid depression and anxiety, this would support the independent liabilities hypothesis rather than the shared liability hypothesis for comorbid depression and anxiety.

## Methods

### Hospital-Based Biobanks

Data for this study were obtained from four different health care system biobanks: Mayo Genome Consortium (MayoGC) linked to the Mayo Clinic in Rochester, Minnesota;^28^ Bio*Me* Biobank linked to the Mount Sinai Health System (MSHS) in New York City, New York;^29^ and two biobanks from the PsycheMERGE (electronic MEdical Record and GEnomics) Network^30^: BioVU linked to Vanderbilt University Medical Center (VUMC) in Nashville, Tennessee;^31^ and the Mass General Brigham (MGB) Biobank linked to the MGB hospital system in Boston, Massachusetts.^32^ Patients enrolled in all four biobanks gave informed consent for use of their EHR data linked to their genetic data. Each site obtained institutional review board approval for the EHR-biobank research.

The EHR includes information on patients such as demographics, medications/prescriptions, laboratory values, billing codes from the International Classification of Diseases, 9^th^ and 10^th^ editions (ICD-9 and ICD-10),^33^ and Current Procedural Terminology (CPT) codes. For the current study, structured EHR data for participants was extracted at the four sites during 2021 and included all patient data before that date (details provided in eMethods).

### Defining cases with depression and anxiety

Depression and anxiety were defined using an initial list of ICD9/10 codes mapped to phecodes (i.e. higher order group of diagnoses, as described and validated by the Phecode map 1.2b1^34^) available from https://phewascatalog.org/phecodes.^35,36^ These definitions were then modified through expert curation from the authors. Depression was defined as having at least one depression-related ICD9/10 code, using an initial list of ICD9/10 codes mapped to the phecode for depression (296.2 and 296.22),^35,36^ with the addition of dysthymic disorder [ICD9:300.4; ICD10:F34.1], depressive type psychosis [ICD9:298.0], and atypical depressive disorder [ICD9:296.82]. Anxiety was defined as having at least one anxiety-related ICD9/10 code, using an initial list of ICD9/10 codes mapped to phecodes for anxiety disorders, generalized anxiety disorders, social phobias and panic disorders, and phobias (300.1, 300.11, 300.12, 300.13, respectively)^35,36^ with the addition of separation anxiety disorder [ICD9: 309.21; ICD10: F93.0] and removing phobic and social anxiety disorder in childhood (ICD10: F93.1 and F93.2) as well as hysteria (ICD9: 300.1) and overanxious disorder (ICD9: 313). The complete list of ICD9/10 codes is presented in eTable 1 and eTable2. As a sensitivity analysis, we made the definitions of depression and anxiety stricter by requiring at least two codes rather than one. Patients with only one diagnostic code were excluded from this sensitivity analysis. In both analyses, controls were patients who had no documented ICD codes for depression and anxiety disorders.

### Genetic data

Samples were genotyped and the genetic data was imputed and processed for quality control at each site as described in the eMethods section. Briefly, DNA samples from blood obtained from study participants were assayed using various genome-wide genotyping arrays. Single nucleotide polymorphisms (SNPs) were excluded using filters for call rate, minor allele frequency (<1%), and Hardy-Weinberg Equilibrium. Individuals were excluded for missingness rate, sex errors, heterozygosity, and relatedness. The study included three ancestries: European, African, Hispanic. MayoGC and MGB included only patients from European ancestries in their analyses due to sample size limitations of other ancestries. Genotype imputation was performed after the initial quality control and converted to best-guess genotypes for all markers with high-quality imputation (dosage-R2 > 0.8). All subsequent analyses were adjusted for principal components (PCs) within each ancestral group to reduce confounding by population substructure.

### Polygenic Risk Scores

To estimate the genetic risk for depression and anxiety, we calculated PRSs using the summary statistics from the Psychiatric Genomic Consortium (PGC) GWAS from Major Depressive Disorder (MDD)^23^ and anxiety (ANX)^22^ working groups, respectively. Both GWASs were performed in European ancestry samples. The PRS calculations were performed separately for each ancestral group at each site. The PRSs were calculated using a pruning-and-thresholding approach implemented by PRSice2^37^ for most sites, and a Bayesian approach implemented by PRS-CS^38^ for the BioVU sample. To avoid the bias of searching for the best p-value threshold to use for PRSice2, we implemented a previously proposed PRS-PCA approach by calculating the PRSs at multiple p-value thresholds (p_T_ = 5e-8, 1e-7, 1e-6, 1e-5, 0.0001, 0.001, 0.01, 0.05, 0.1, 0.2, 1), performing a PC-analysis on the resulting set of PRSs, and keeping the first PC.^38,39^ This resulted in one PRS for depression and one PRS for anxiety for every individual in each of the samples.

### Statistical Analysis

All analyses were stratified by site and ancestral group. Logistic regression was first used to assess whether each PRS (depression or anxiety) was associated with the matching EHR-defined diagnosis while adjusting for PCs. Next, we used multinomial regression to assess how well the combination of the MDD-PRS and ANX-PRS jointly predict the comorbidity of depression and anxiety by including both MDD-PRS and ANX-PRS in the same model. The categorical outcome used in the multinomial model classified patients into one of four different groups: neither depression or anxiety (controls), depression-only, anxiety-only, and both. Effect sizes for PRS analyses were represented by odds ratios and percent of variation explained on the liability scale (assuming a 20% prevalence of both depression and anxiety in the general population).

## Results

Figure 1 describes the distribution of depression and anxiety diagnoses defined from the EHR (1+ codes) at each site (Total N = 140,947). Around 70% of each biobank sample had received no ICD codes for depression or anxiety with the exception of MGB which had a higher prevalence of anxiety-related codes (35%) and thus a lower proportion of controls. When comparing diagnostic rates across self-reported race and ethnicity in Bio*Me*, African American and Hispanic participants had higher rates of depression codes (22% and 28%, respectively) than self-identified White participants (15%), while White participants from Bio*Me* as well as BioVU had slightly higher rates of anxiety-related codes than African American and Hispanic participants (∼8% vs. ∼6%, respectively). The full demographic characteristics and EHR summaries by site are shown in eTable 3.

**Figure 1.**
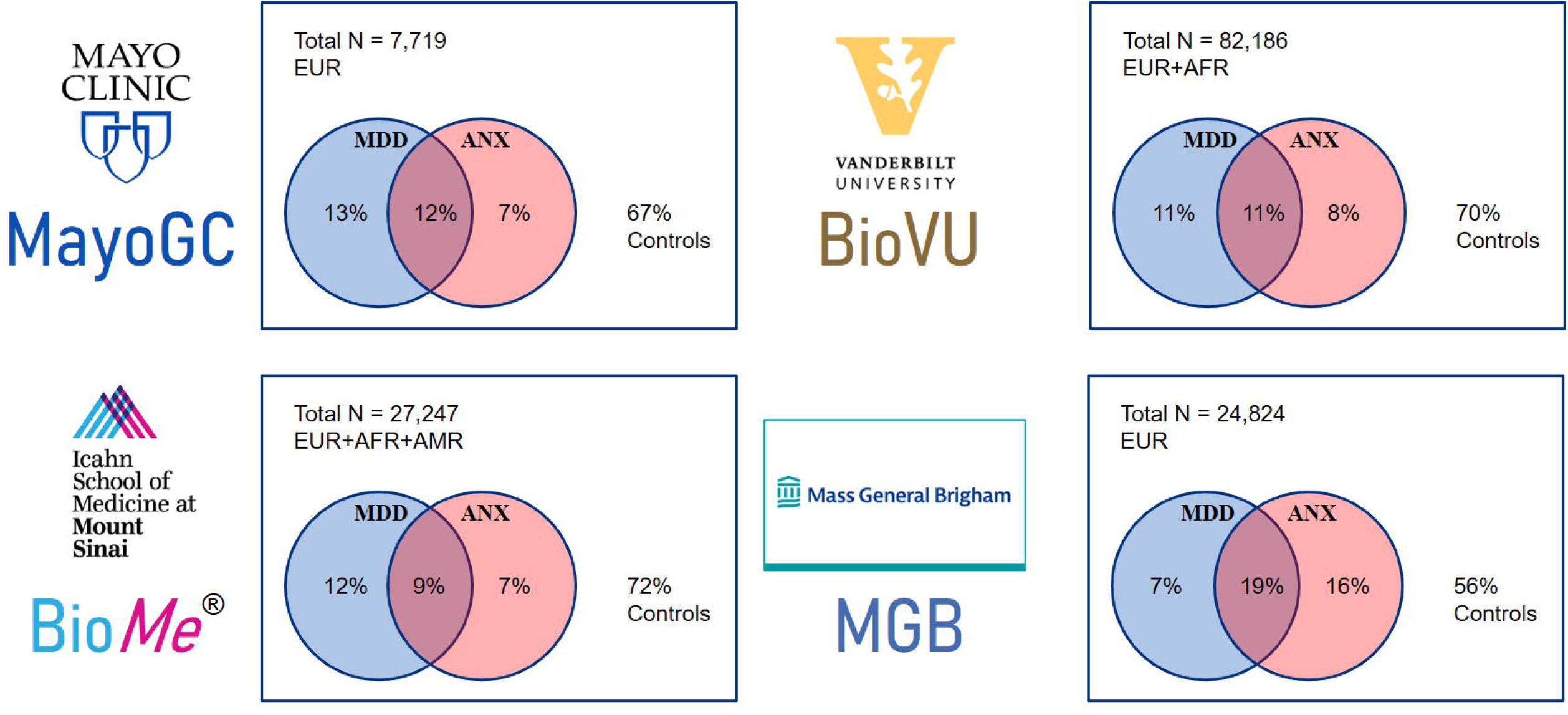
Description of each biobank. Distribution of depression (MDD) and anxiety (ANX) diagnoses at each site specified by at least one diagnosis code from the EHR. Each site’s sample size and ancestries (EUR=European, AFR=African/African American, AMR=Hispanic) are included in the top left of each site plot.

### PRS prediction of depression and anxiety separately

We first tested whether the MDD-PRS and ANX-PRS, trained on samples from European ancestries that are largely clinically ascertained, were associated with depression and anxiety diagnoses (1 or more ICD codes) in the EHR, respectively. Figure 2 shows the PRS predictive performance for each site and ancestry group as measured by the percent variation explained on the liability scale. Among those of European ancestry, the MDD-PRS explained a significant proportion of variation in the liability of depression (0.5-1.2% across sites). While the MDD-PRS was also associated with depression in the African American and Hispanic participants from BioVU and Bio*Me*, the predictive performance was markedly attenuated, explaining only 0.11% to 0.35% of variation in the liability of depression. When requiring two or more ICD codes, the percent of variation explained increased slightly in almost all cases with the exception of the African American sample of Bio*Me* (R^2^=0.12%; p-value = 0.07).

**Figure 2.**
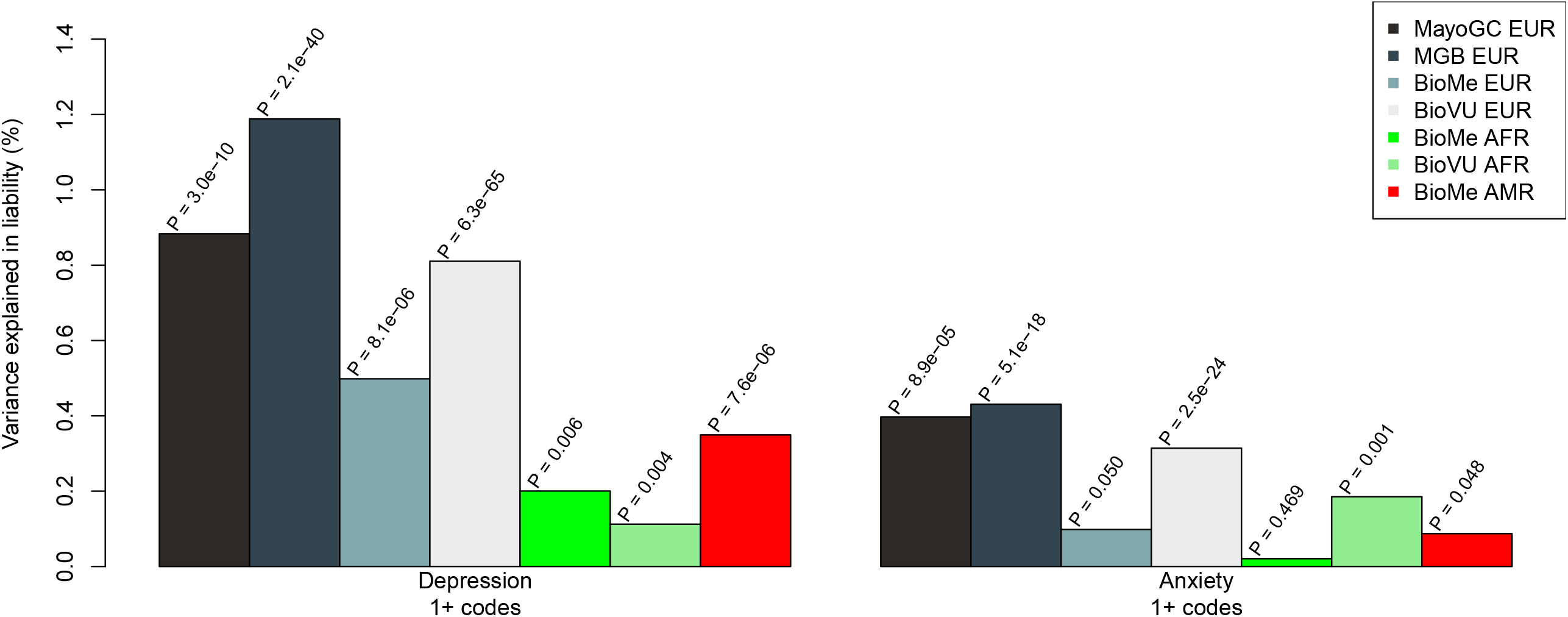
PRS prediction of depression and anxiety separately. Site- and ancestry-specific association of MDD-PRS and ANX-PRS with MDD and ANX, respectively, defined by having at least one ICD code from the EHR. Performance is measured by variance explained by the PRS on the liability scale (assuming 20% population prevalence for both disorders). P-values for each association are listed above each bar.

In comparison to the performance of the MDD-PRS, the ANX-PRS explained less variation in the liability of anxiety (0.1-0.43% across sites) and was only a marginally significant predictor in Bio*Me* (R^2^ = 0.1%; p-value = 0.0504). Similar to MDD-PRS, the performance of the ANX-PRS was reduced in the non-EUR cohorts and only significant in the BioVU African American sample (R^2^ = 0.19%; p-value = 0.001). The full results are shown in eTable 4.

### Joint PRS prediction of depression and anxiety comorbidity

Next, we tested whether both PRSs were associated with the comorbidity of depression and anxiety using a multinomial outcome specifying whether a participant had neither diagnosis (N=95,992), depression-only (N=14,918), anxiety-only (N=12,682), or comorbid (N=17,355). In the combined meta-analysis across participants of European ancestry at each site (Total N=109,592) as shown in Table 1, both MDD-PRS and ANX-PRS were significantly associated with each case subgroup compared to controls after adjusting for one another. Notably, both the MDD-PRS (OR=1.20 per SD increase in PRS; 95% CI= 1.18-1.23) and the ANX-PRS (OR=1.07; 95% CI=1.05-1.09) had the largest effect size in the comorbid group when compared to controls. Furthermore, the 95% confidence interval for the MDD-PRS effect size did not overlap across groups showing a gradient of genetic risk across the isolated conditions and the comorbidity.

**Table 1.**
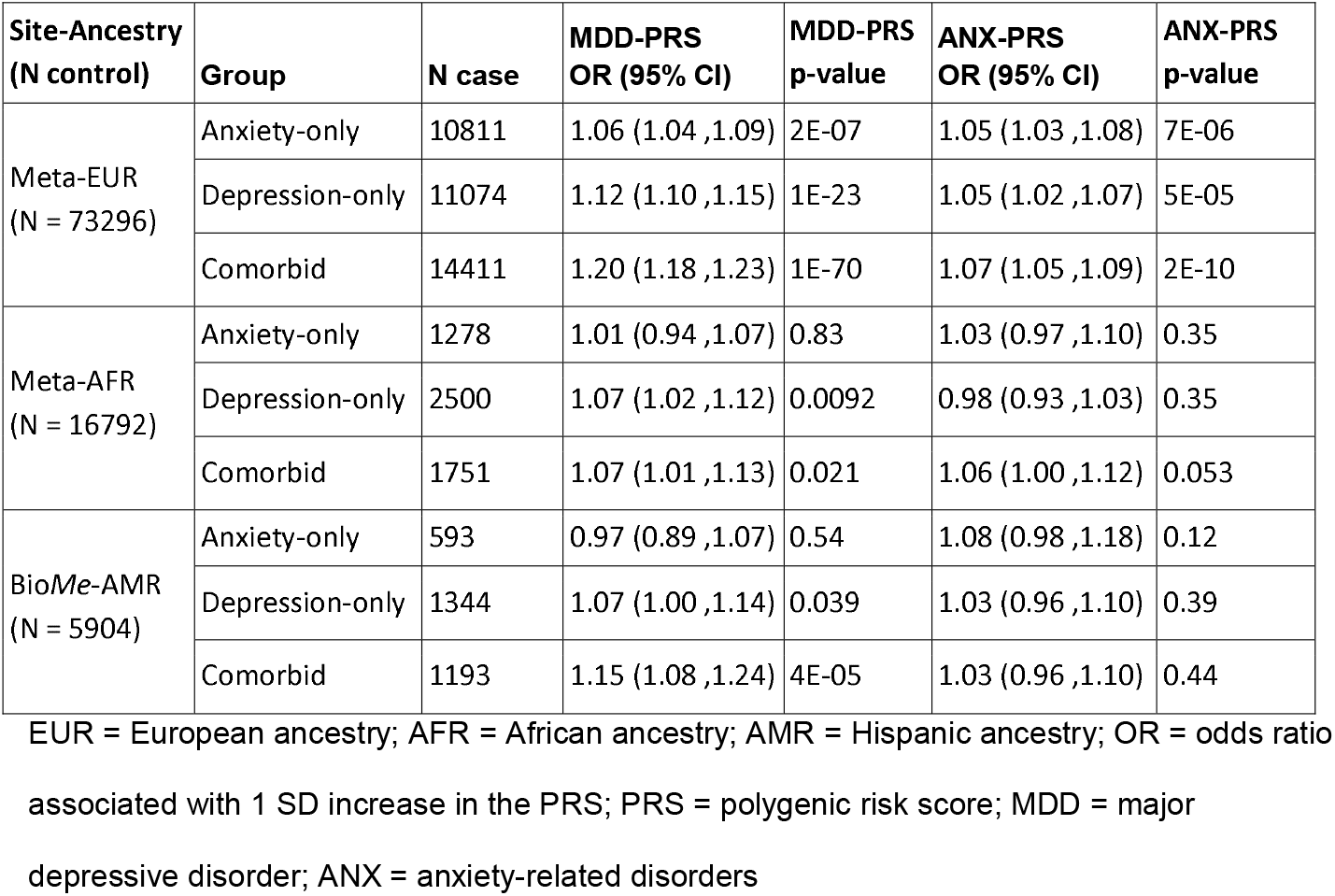
Joint PRS prediction of depression and anxiety comorbidity from multinomial model.

In the meta-analysis of African American participants from Bio*Me* and BioVU (N=22,321), after adjusting for the contribution of the ANX-PRS, the MDD-PRS was associated only with depression-only (N=2,500; OR=1.07; 95% CI=1.02-1.12) and comorbid (N=1,751; OR=1.07; 95% CI=1.01-1.13) groups when compared to controls (N=16,792) and not for ANX-only (N=1,278; OR=1.01; 95% CI=0.94-1.07). Among the Hispanic participants from Bio*Me* (N=9,034), the MDD-PRS was only predictive of the comorbid group (N=1,193; OR=1.15; 95% CI=1.08-1.24) compared to controls (N=5,904). After adjusting for the contribution of MDD-PRS, the ANX-PRS was not predictive of any case subgroup compared to controls among African American and Hispanic participants. When requiring two or more ICD codes, the results did not substantially change. The full results by ancestry and by site are presented in eTable5.

## Discussion

Prior GWASs of depression and anxiety disorders typically have not considered the common comorbidity between depression and anxiety. Therefore, little is known about how genetic risk impacts the comorbidity. To study this, we performed a multi-site and multi-ancestry PRS analysis across four different EHR-linked biobanks using genetic risk of depression and anxiety. The MDD-PRS predicted diagnosis of depression in the EHR in all ancestries and at all sites.

However, ANX-PRS was less predictive of anxiety diagnoses, with the ANX-PRS having a smaller effect size than MDD-PRS and was not significantly associated with anxiety disorders among all ancestries in Bio*Me*. Importantly though, in our model of the comorbidity in European ancestry samples, the MDD-PRS showed a gradient of risk with respect to diagnosis of depression and anxiety such that patients with the comorbidity had the highest MDD-PRS followed by those with depression-only, anxiety-only, and neither diagnosis.

This observed gradient of depression genetic risk across the comorbidity could be informative from a clinical perspective. While depression and anxiety have a genetic correlation close to one, the MDD-PRS was higher for patients with depression-only than those with anxiety-only after adjusting for ANX-PRS implying that the additive genetic factors differ between the two disorders. Further, we found that the comorbid group was at the highest level of genetic risk for depression, which could indicate that genetic factors play a larger role in those with the comorbidity compared with only one diagnosed disorder.

However, it is important to consider how our study design could also impact these results. In our study, depression and anxiety were defined with ICD codes that have only modest concordance with cases identified by structured or semi-structured research assessments.^40^ Thus, while our finding that MDD+ANX has the highest genetic risk of MDD could reflect true biological differences, it might instead reflect the fact that the MDD+ANX comorbidity is more clinically conspicuous and therefore better captured in the EHR because it is associated with more severe depressive symptoms, higher levels of impairment, poorer clinical outcomes, and more suicidal ideation.^3–6^ These patients may be more likely to be referred for specialty care resulting in a higher likelihood of a depression or anxiety diagnosis. If so, our PRS analysis may then still reflect biologically determined greater symptom severity. Consequently, repeating this study in a large sample with more rigorous diagnostic assessments would be of value.

A strength of this study is that a larger range of the phenome can be easily and cost-effectively captured through the EHR. While non-EHR clinical studies may have more in-depth assessment of the phenotype of interest, they typically lack comprehensive assessment of important comorbidities. These types of studies also are not feasible for large samples such as biobanks. It is also important to acknowledge the limitations of EHR-based studies, including this study, reliance on ICD codes from the EHR can result in misclassification of cases and controls. To explore the impact of misclassification of cases, we performed a sensitivity analysis restricting cases to those with two or more ICD depression or anxiety codes and found no large deviations from the primary results based on one or more codes. A strategy to improve case/control ascertainment from the EHR is to use natural language processing (NLP) to incorporate clinical notes into the classification system.^41^ Such an approach requires NLP algorithms that translate well across health systems, which is an active area of research. Heterogeneity of diagnosis prevalence between the different sites may have also contributed to differential power in the analysis. Given this, it is perhaps more striking that the PRSs demonstrated significant association despite heterogeneity of patient populations, diagnostic practices, and differences in EHR systems across sites, albeit with small amounts of variance explained, for diagnosis of depression and anxiety.

Finally, this study of depression and anxiety was one of the first to include samples from diverse groups. However, the sample sizes of African and Hispanic ancestry patients were much smaller than the European ancestry group, resulting in lower power to detect associations with genetic risk. Furthermore, the constructed PRSs were based on cohorts with European ancestry, which are known to have poorer prediction in non-European ancestries.^42^ The African American and Hispanic patients also had higher rates of depression diagnoses which is contrary to what might be expected based on prior epidemiological^43^ or clinical^44^ research. Moreover, diagnostic biases^45^ and decreased healthcare access for people from marginalized communities^46^ can contribute to the misclassification in these groups and thus further decrease in power.

Our study found that genetic risks of depression and anxiety are jointly informative of depression and anxiety with stronger associations with depression diagnosis. Our observation that genetic vulnerabilities to depression and to a lesser extent anxiety make distinct contributions to risk of comorbid depression and anxiety challenges strict interpretations of the shared liability hypothesis. It also argues against ICD-10-CM mixed anxiety and depressive disorder, which has no correlate in DSM-5,^47^ as a distinct nosological entity.

In addition to needing to follow up this analysis in a large sample with research diagnostic assessments, future collections should focus on inclusion of patients of non-European ancestries in order to improve PRS transferability.^42^ Given our findings that the current PRSs can differentiate between those with depression-only, anxiety-only, and the comorbidity, GWAS of each of these groups in large samples could be informative for future PRS analyses. Even without these GWAS to create comorbid-specific PRS, we found that the MDD-PRS and ANX-PRS were independently associated with the comorbidity of depression and anxiety in the EHR. This supports the hypothesis that the correlated disorders represent distinct nosological entities.

## Data Availability

Data in the present study is available through application to each biobank and is unable to be shared otherwise.

## Acknowledgements

This study was supported by the National Institute of Mental Health (NIMH) grants: R01MH121924, R01MH121923, R01MH121922, and R01MH121921. KWC was supported in part by funding from the NIMH (K08MH127413) and a NARSAD Brain and Behavior Foundation Young Investigator Award. YNG was supported in part by a Moynihan Clinical Research Fellowship from the Leon Levy Foundation and Award Number R25MH086466 from the NIMH. LKD and JWS were supported by R01MH118233. LKD is also supported by R56MH120736. Vanderbilt EHR resources were supported by the National Center for Research Resources, Grant UL1 RR024975-01, and is now at the National Center for Advancing Translational Sciences, Grant 2 UL1 TR000445-06. The content is solely the responsibility of the authors and does not necessarily represent the official views of the NIH. Vanderbilt University Medical Center’s BioVU is supported by numerous sources: institutional funding, private agencies, and federal grants. These include the NIH funded Shared Instrumentation Grant S10RR025141; and CTSA grants UL1TR002243, UL1TR000445, and UL1RR024975. Genomic data are also supported by investigator-led projects that include U01HG004798, R01NS032830, RC2GM092618, P50GM115305, U01HG006378, U19HL065962, R01HD074711; and additional funding sources listed at https://victr.vumc.org/biovu-funding/.

## References

1. Global Burden of Disease Study 2013 Collaborators. Global, regional, and national incidence, prevalence, and years lived with disability for 301 acute and chronic diseases and injuries in 188 countries, 1990-2013: a systematic analysis for the Global Burden of Disease Study 2013. Lancet. 2015;386(9995):743–800.

2. Kessler RC, Gruber M, Hettema JM, Hwang I, Sampson N, Yonkers KA. Co-morbid major depression and generalized anxiety disorders in the National Comorbidity Survey follow-up. Psychol Med. 2008;38(3):365–374.

3. Nam YY, Kim CH, Roh D. Comorbid panic disorder as an independent risk factor for suicide attempts in depressed outpatients. Compr Psychiatry. 2016;67:13–18.

4. Pollack MH. Comorbid anxiety and depression. J Clin Psychiatry. 2005;66 Suppl 8:22–29.

5. Joffe RT, Bagby RM, Levitt A. Anxious and nonanxious depression. Am J Psychiatry. 1993;150(8):1257–1258.

6. Zhou Y, Cao Z, Yang M, et al. Comorbid generalized anxiety disorder and its association with quality of life in patients with major depressive disorder. Scientific Reports. 2017;7(1). doi:10.1038/srep40511

7. Blanco C, Krueger RF, Hasin DS, et al. Mapping Common Psychiatric Disorders. JAMA Psychiatry. 2013;70(2):199. doi:10.1001/jamapsychiatry.2013.281

8. Krueger RF. The Structure of Common Mental Disorders. Archives of General Psychiatry. 1999;56(10):921. doi:10.1001/archpsyc.56.10.921

9. Kotov R, Krueger RF, Watson D, et al. The Hierarchical Taxonomy of Psychopathology (HiTOP): A dimensional alternative to traditional nosologies. J Abnorm Psychol. 2017;126(4):454–477.

10. American Psychiatric Association. Diagnostic and Statistical Manual of Mental Disorders, Fifth Edition, Text Revision (DSM-5-TR(tm)).; 2022.

11. Ressler KJ, Nemeroff CB. Role of serotonergic and noradrenergic systems in the pathophysiology of depression and anxiety disorders. Depress Anxiety. 2000;12 Suppl 1:2–19.

12. Zorn JV, Schür RR, Boks MP, Kahn RS, Joëls M, Vinkers CH. Cortisol stress reactivity across psychiatric disorders: A systematic review and meta-analysis. Psychoneuroendocrinology. 2017;77:25–36.

13. Nutt D. Anxiety and depression: individual entities or two sides of the same coin? Int J Psychiatry Clin Pract. 2004;8 Suppl 1:19–24.

14. Craske MG. Transdiagnostic treatment for anxiety and depression. Depress Anxiety. 2012;29(9):749–753.

15. Blanco C, Wall MM, Feng T, Olfson M. Evaluating the modified common liability hypothesis of psychiatric comorbidity. J Psychiatr Res. 2021;141:9–15.

16. Greenebaum SLA, Nierenberg AA. More on the NIMH RDoC initiative. Bipolar Disorders. 2020;22(1):11–12. doi:10.1111/bdi.12874

17. Dillon DG, Rosso IM, Pechtel P, Killgore WDS, Rauch SL, Pizzagalli DA. Peril and pleasure: an rdoc-inspired examination of threat responses and reward processing in anxiety and depression. Depress Anxiety. 2014;31(3):233–249.

18. Takahashi Y, Yamagata S, Ritchie SJ, Barker ED, Ando J. Etiological pathways of depressive and anxiety symptoms linked to personality traits: A genetically-informative longitudinal study. J Affect Disord. 2021;291:261–269.

19. de Graaf R, Bijl RV, Smit F, Vollebergh WAM, Spijker J. Risk factors for 12-month comorbidity of mood, anxiety, and substance use disorders: findings from the Netherlands Mental Health Survey and Incidence Study. Am J Psychiatry. 2002;159(4):620–629.

20. Wray NR, Ripke S, Mattheisen M, et al. Genome-wide association analyses identify 44 risk variants and refine the genetic architecture of major depression. Nat Genet. 2018;50(5):668–681.

21. Levey DF, Stein MB, Wendt FR, et al. Bi-ancestral depression GWAS in the Million Veteran Program and meta-analysis in >1.2 million individuals highlight new therapeutic directions. Nat Neurosci. 2021;24(7):954–963.

22. Purves KL, Coleman JRI, Meier SM, et al. A major role for common genetic variation in anxiety disorders. Mol Psychiatry. 2020;25(12):3292–3303.

23. Howard DM, Adams MJ, Clarke TK, et al. Genome-wide meta-analysis of depression identifies 102 independent variants and highlights the importance of the prefrontal brain regions. Nat Neurosci. 2019;22(3):343–352.

24. Giannakopoulou O, Lin K, Meng X, et al. The Genetic Architecture of Depression in Individuals of East Asian Ancestry: A Genome-Wide Association Study. JAMA Psychiatry. 2021;78(11):1258–1269.

25. Levey DF, Gelernter J, Polimanti R, et al. Reproducible Genetic Risk Loci for Anxiety: Results From 200,000 Participants in the Million Veteran Program. American Journal of Psychiatry. 2020;177(3):223–232. doi:10.1176/appi.ajp.2019.19030256

26. Peterson RE, Kuchenbaecker K, Walters RK, et al. Genome-wide Association Studies in Ancestrally Diverse Populations: Opportunities, Methods, Pitfalls, and Recommendations. Cell. 2019;179(3):589–603.

27. Zheutlin AB, Dennis J, Karlsson Linnér R, et al. Penetrance and Pleiotropy of Polygenic Risk Scores for Schizophrenia in 106,160 Patients Across Four Health Care Systems. Am J Psychiatry. 2019;176(10):846–855.

28. Bielinski SJ, Chai HS, Pathak J, et al. Mayo Genome Consortia: a genotype-phenotype resource for genome-wide association studies with an application to the analysis of circulating bilirubin levels. Mayo Clin Proc. 2011;86(7):606–614.

29. Belbin GM, Odgis J, Sorokin EP, et al. Genetic identification of a common collagen disease in puerto ricans via identity-by-descent mapping in a health system. Elife. 2017;6. doi:10.7554/eLife.25060

30. Roden DM, Pulley JM, Basford MA, et al. Development of a large-scale de-identified DNA biobank to enable personalized medicine. Clin Pharmacol Ther. 2008;84(3):362–369.

31. Karlson E, Boutin N, Hoffnagle A, Allen N. Building the Partners HealthCare Biobank at Partners Personalized Medicine: Informed Consent, Return of Research Results, Recruitment Lessons and Operational Considerations. Journal of Personalized Medicine. 2016;6(1):2. doi:10.3390/jpm6010002

32. WHO, World Health Organization. The ICD-10 Classification of Mental and Behavioural Disorders: Diagnostic Criteria for Research. World Health Organization; 1993.

33. Denny JC, Ritchie MD, Basford MA, et al. PheWAS: demonstrating the feasibility of a phenome-wide scan to discover gene-disease associations. Bioinformatics. 2010;26(9):1205–1210.

34. Wei WQ, Bastarache LA, Carroll RJ, et al. Evaluating phecodes, clinical classification software, and ICD-9-CM codes for phenome-wide association studies in the electronic health record. PLoS One. 2017;12(7):e0175508.

35. He M, Santiago Ortiz AJ, Marshall J, et al. PBI40 ICD-9 TO ICD-10 MAPPING FOR RESEARCH IN BIOLOGICS AND BIOSIMILARS USING ADMINISTRATIVE HEALTHCARE DATA. Value in Health. 2019;22:S55. doi:10.1016/j.jval.2019.04.122

36. Choi SW, O’Reilly PF. PRSice-2: Polygenic Risk Score software for biobank-scale data. Gigascience. 2019;8(7). doi:10.1093/gigascience/giz082

37. Ge T, Chen CY, Ni Y, Feng YCA, Smoller JW. Polygenic prediction via Bayesian regression and continuous shrinkage priors. Nat Commun. 2019;10(1):1776.

38. Coombes BJ, Ploner A, Bergen SE, Biernacka JM. A principal component approach to improve association testing with polygenic risk scores. Genet Epidemiol. 2020;44(7):676–686.

39. Fiest KM, Jette N, Quan H, et al. Systematic review and assessment of validated case definitions for depression in administrative data. BMC Psychiatry. 2014;14:289.

40. Ford E, Carroll JA, Smith HE, Scott D, Cassell JA. Extracting information from the text of electronic medical records to improve case detection: a systematic review. Journal of the American Medical Informatics Association. 2016;23(5):1007–1015. doi:10.1093/jamia/ocv180

41. Martin AR, Kanai M, Kamatani Y, Okada Y, Neale BM, Daly MJ. Current clinical use of polygenic scores will risk exacerbating health disparities. doi:10.1101/441261

42. Breslau J, Kendler KS, Su M, Gaxiola-Aguilar S, Kessler RC. Lifetime risk and persistence of psychiatric disorders across ethnic groups in the United States. Psychol Med. 2005;35(3):317–327.

43. Stockdale SE, Lagomasino IT, Siddique J, McGuire T, Miranda J. Racial and ethnic disparities in detection and treatment of depression and anxiety among psychiatric and primary health care visits, 1995-2005. Med Care. 2008;46(7):668–677.

44. Gara MA, Vega WA, Arndt S, et al. Influence of patient race and ethnicity on clinical assessment in patients with affective disorders. Arch Gen Psychiatry. 2012;69(6):593–600.

45. Bailey RK, Mokonogho J, Kumar A. Racial and ethnic differences in depression: current perspectives. Neuropsychiatr Dis Treat. 2019;15:603–609.

46. Möller HJ, Bandelow B, Volz HP, Barnikol UB, Seifritz E, Kasper S. The relevance of “mixed anxiety and depression” as a diagnostic category in clinical practice. Eur Arch Psychiatry Clin Neurosci. 2016;266(8):725–736.

