## Supplementary Material for "The contribution of genetic risk to the comorbidity of depression and anxiety: a multi-site electronic health records study"

**eMethods**

### **Cohort Descriptions**

#### ***The Mayo Genome Consortia (MayoGC)***

The MayoGC includes participants, mostly control samples, from 10 different genetic studies that had recruited patients from the Mayo Clinic Health System^1^. Participants’ DNA isolated from whole blood was genotyped using different genotyping platforms, and each genotyping batch was run through the same genotype quality control (QC) pipeline (Below Table). In this QC pipeline, SNPs were excluded using filters for call rate (<95%), minor allele frequency (<1%), and Hardy-Weinberg Equilibrium. Individuals were excluded for excessive missing data (>5%), sex errors, abnormal heterozygosity, and relatedness (removing a random individual from any pair with kinship coefficient > 0.2). After QC, genotype data from each study was imputed using the Michigan Imputation server (Minimac4 1.2.1) with the HRC reference panel (Reference Panel: apps@hrc-r1.1 [hg19])^2^. Imputed genotypes were converted to best-guess genotypes for all markers with high-quality imputation (dosage-R2 > 0.8). Cases and controls were ascertained based on EHR data from 7719 Mayo Clinic Health System patients from the MayoGC^1^, using EHR data extracted on May 5, 2021, which included all diagnoses before that date. The Institutional Review Board of Mayo Clinic approved this study.

| **Study Name** | **N** | **Genotyping Array** |
| --- | --- | --- |
| MayoGC-BRAIN | 142 | Illumina 610K |
| MayoGC-Cerhan3 | 321 | Illumina Omni Express |
| MayoGC-CLL | 199 | Affymetrix 6.0 |
| MayoGC-EM | 2910 | Illumina 660K |
| MayoGC-Lung | 138 / 127 | Illumina 370K / Illumina 610K |
| MayoGC-NHL | 169 | Illumina 660K |
| MayoGC-Ovarian1 | 432 | Illumina 610K |
| MayoGC-Ovarian2 | 409 | Human Omni 2.5-4v1 |
| MayoGC-PA | 287 / 232 | PanScan I-550K / PanScan II-610K |
| MayoGC-VTE | 1891 | Illumina 660K |

#### ***BioMe***

#### Cases and controls were ascertained using EHR data from 26,218 patients from the Bio*Me* biobank in Mount Sinai Health System^1^. EHR data for the participants was extracted in September 2020 and included any diagnostics on or before that date. DNA for GWAS analysis was isolated from whole blood and genotyped using the Illumina Global Screening Array (GSA) platform. The Institutional Review Board approved this study. QC and imputation have been described previously.^3^ Briefly, in the QC pipeline, SNPs were excluded using filters for call rate (<95%), minor allele frequency (<1%), and Hardy-Weinberg Equilibrium (p < 1e-5). Individuals were excluded for excessive missing data (>5%), sex errors, abnormal heterozygosity (±6 SD) and relatedness (removing a random individual from any pair with kinship coefficient > 0.2). Imputation was performed on phased haplotypes using IMPUTE(v2.3.2)^4^ with the phase III Thousand Genomes data as reference panel, and the addition of the following flag: ‘‘-filt_rules_1 ‘ALL<0.0002’ ‘ALL>0.9998’.

#### ***BioVU***

#### Cases and controls were ascertained using EHR data from 82186 patients from the Vanderbilt University Medical Center (VUMC)^5^. EHR data for the participants was extracted in June 2021 and included any diagnostics on or before that date. DNA for GWAS analysis was isolated from whole blood and genotyped using the Illumina MEGAEX platform. The VUMC Institutional Review Board approved this study. In the QC pipeline, SNPs were excluded using filters for call rate (<2%), minor allele frequency (<1%), and Hardy-Weinberg Equilibrium (p < 5e-5). Individuals were excluded for excessive missing data (>2%), sex errors, abnormal heterozygosity (|Fhet| > 0.2) and relatedness (removing a random individual from any pair with kinship coefficient > 0.2). After QC, genotypes were imputed using SHAPEIT^6^/IMPUTE4^4^ with the 1000 genomes phase I reference panel. Genotype imputation was performed after the initial QC and converted to best-guess genotypes for all markers with high-quality imputation (INFO > 0.3).

#### ***Partners Biobank (MGB)***

Cases and controls were ascertained using EHR data from 24,842 patients from the Mass General Brigham (MGB) Health System. EHR data for the participants was extracted in February 2020 and included any diagnoses on or before that date. DNA for GWAS analysis was isolated from blood and genotyped using Illumina arrays (MEGA, MEGAEX, and MEG BeadChip). The Institutional Review Board approved this study. In the QC pipeline, SNPs were excluded using filters for call rate (<95%), and Hardy-Weinberg Equilibrium (p<1e-10). Individuals were excluded for excessive missing data (>2%), sex errors, abnormal heterozygosity (±3 SD) and relatedness (removing a random individual from any pair with kinship coefficient > 0.2). After QC, batches from genotyping arrays were merged and then imputed using the Michigan Imputation server (Minimac4 1.2.1) with the HRC/1KG reference panel (Reference Panel: apps@hrc-r1.1 [hg19])^2^. Genotype imputation was performed after the initial QC and converted to best-guess genotypes for all markers with high-quality imputation (dosage-R2 > 0.8) and common minor allele frequency (>1%).

##### **eFigure 1.** **Sensitivity analysis for PRS prediction of depression and anxiety separately.** Site- and ancestry-specific association of MDD-PRS and ANX-PRS with MDD and ANX, respectively, defined by having at least two ICD codes from the EHR. Performance is measured by variance explained by the PRS on the liability scale (assuming 20% population prevalence for both disorders). P-values for each association are listed above each bar.

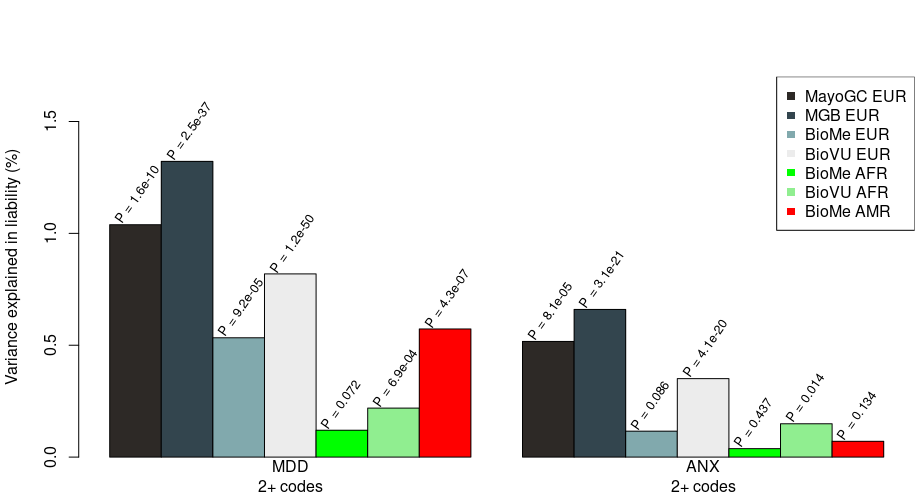

### **eTable 1. ICD 9/10 codes used for defining depression.**

| **ICD9/10** | **ICD** | **Phecode** | **Description** |
| --- | --- | --- | --- |
| 296.2 | ICD9 | 296.22 | Major depressive disorder, single episode |
| 296.2 | ICD9 | 296.22 | Major depressive disorder, single episode, unspecified degree |
| 296.21 | ICD9 | 296.2 | Major depressive disorder, single episode, mild degree |
| 296.22 | ICD9 | 296.22 | Major depressive disorder, single episode, moderate degree |
| 296.23 | ICD9 | 296.22 | Major depressive disorder, single episode, severe degree, w/o psychotic behavior |
| 296.24 | ICD9 | 296.22 | Major depressive disorder, single episode, severe degree, specified psychotic behavior |
| 296.25 | ICD9 | 296.22 | Major depressive disorder, single episode, in partial or unspecified remission |
| 296.26 | ICD9 | 296.22 | Major depressive disorder, single episode in full remission |
| 296.3 | ICD9 | 296.22 | Major depressive disorder, recurrent episode |
| 296.3 | ICD9 | 296.22 | Major depressive disorder, recurrent episode, unspecified degree |
| 296.31 | ICD9 | 296.2 | Major depressive disorder, recurrent episode, mild degree |
| 296.32 | ICD9 | 296.22 | Major depressive disorder, recurrent episode, moderate degree |
| 296.33 | ICD9 | 296.22 | Major depressive disorder, recurrent episode, severe degree, w/o psychotic behavior |
| 296.34 | ICD9 | 296.22 | Major depressive disorder, recurrent episode, severe degree, spec. psychotic behavior |
| 296.35 | ICD9 | 296.22 | Major depressive disorder, recurrent episode, in partial or unspecified remission |
| 296.36 | ICD9 | 296.22 | Major depressive disorder, recurrent episode, in full remission |
| 311 | ICD9 | 296.2 | Depressive disorder NEC |
| 300.4 | ICD9 | 300.4 | Dysthymic disorder |
| 298.0 | ICD9 | 295.3 | Depressive type psychosis |
| 296.82 | ICD9 | 296.1 | Atypical depressive disorder |
| F32 | ICD10 | 296.22 | Major depressive disorder, single episode |
| F32.0 | ICD10 | 296.22 | Major depressive disorder, single episode, mild |
| F32.1 | ICD10 | 296.22 | Major depressive disorder, single episode, moderate |
| F32.2 | ICD10 | 296.22 | Major depressive disorder, single episode, severe without psychotic features |
| F32.3 | ICD10 | 296.22 | Major depressive disorder, single episode, severe with psychotic features |
| F32.4 | ICD10 | 296.22 | Major depressive disorder, single episode, in partial remission |
| F32.5 | ICD10 | 296.22 | Major depressive disorder, single episode, in full remission |
| F32.8 | ICD10 | 296.22 | Other depressive episodes |
| F32.89 | ICD10 | 296.22 | Single episode of 'masked' depression NOS |
| F32.9 | ICD10 | 296.22 | Major depressive disorder, single episode, unspecified |
| F33 | ICD10 | 296.22 | Major depressive disorder, recurrent |
| F33.0 | ICD10 | 296.22 | Major depressive disorder, recurrent, mild |
| F33.1 | ICD10 | 296.22 | Major depressive disorder, recurrent, moderate |
| F33.2 | ICD10 | 296.22 | Major depressive disorder, recurrent severe without psychotic features |
| F33.3 | ICD10 | 296.22 | Major depressive disorder, recurrent, severe with psychotic symptoms |
| F33.4 | ICD10 | 296.22 | Major depressive disorder, recurrent, in remission |
| F33.40 | ICD10 | 296.22 | Major depressive disorder, recurrent, in remission, unspecified |
| F33.41 | ICD10 | 296.22 | Major depressive disorder, recurrent, in partial remission |
| F33.42 | ICD10 | 296.22 | Major depressive disorder, recurrent, in full remission |
| F33.8 | ICD10 | 296.22 | Other recurrent depressive disorders |
| F33.9 | ICD10 | 296.22 | Major depressive disorder, recurrent, unspecified |
| F34.1 | ICD10 | 300.4 | Dysthymic disorder |

### **eTable 2. ICD 9/10 codes used for defining anxiety.**

| **ICD9/10** | **ICD** | **PheCode** | **ICD9 String** |
| --- | --- | --- | --- |
| 293.84 | ICD9 | 300.1 | Anxiety disorder in conditions classified elsewhere |
| 300 | ICD9 | 300.1 | Anxiety states |
| 300 | ICD9 | 300.1 | Anxiety state unspecified |
| 300.01 | ICD9 | 300.12 | Panic disorder without agoraphobia |
| 300.02 | ICD9 | 300.11 | Generalized anxiety disorder |
| 300.09 | ICD9 | 300.1 | Other anxiety states |
| 300.2 | ICD9 | 300.13 | Phobia unspecified |
| 300.21 | ICD9 | 300.12 | Agoraphobia with panic disorder |
| 300.22 | ICD9 | 300.12 | Agoraphobia without mention of panic attacks |
| 300.23 | ICD9 | 300.12 | Social phobia |
| 300.29 | ICD9 | 300.13 | Other isolated or specific phobias |
| 309.21 | ICD9 | 313 | Separation anxiety disorder |
| F06.4 | ICD10 | 300.1 | Organic anxiety disorder |
| F40 | ICD10 | 300.13 | Phobic anxiety disorders |
| F40.0 | ICD10 | 300.12 | Agoraphobia |
| F40.1 | ICD10 | 300.12 | Social phobias |
| F40.2 | ICD10 | 300.13 | Specific (isolated) phobias |
| F40.8 | ICD10 | 300.13 | Other phobic anxiety disorders |
| F40.9 | ICD10 | 300.13 | Phobic anxiety disorder, unspecified |
| F41.0 | ICD10 | 300.12 | Panic disorder [episodic paroxysmal anxiety] |
| F41.1 | ICD10 | 300.11 | Generalized anxiety disorder |
| F41.2 | ICD10 | 300.1 | Mixed anxiety and depressive disorder |
| F41.3 | ICD10 | 300.1 | Other mixed anxiety disorders |
| F41.8 | ICD10 | 300.1 | Other specified anxiety disorders |
| F41.9 | ICD10 | 300.1 | Anxiety disorder, unspecified |

**eTable 3. Demographic and EHR summary statistics across site.**

|  | **Site** | **MayoGC** | **MGB** | **Bio*Me*** | **Bio*Me*** | **Bio*Me*** | **BioVU** | **BioVU** |
| --- | --- | --- | --- | --- | --- | --- | --- | --- |
|  | **Ancestry** | **EUR** | **EUR** | **EUR** | **AFR** | **AMR** | **EUR** | **AFR** |
| **Variable** | **N** | 7719 | 24824 | 10146 | 7038 | 9034 | 66903 | 15283 |
|  | Control | 5216 (67.6%) | 14208 (57.2%) | 7800 (76.9%) | 5058 (71.9%) | 5904 (65.4%) | 46072 (68.9%) | 11734 (76.8%) |
| Diagnosis group (1+ Code) | MDD-only | 994 (12.9%) | 1922 (7.7%) | 857 (8.4%) | 1028 (14.6%) | 1344 (14.9%) | 7301 (10.9%) | 1472 (9.6%) |
|  | ANX-only | 556 (7.2%) | 3869 (15.6%) | 814 (8%) | 405 (5.8%) | 593 (6.6%) | 5572 (8.3%) | 873 (5.7%) |
|  | MDD+ANX | 953 (12.3%) | 4825 (19.4%) | 675 (6.7%) | 547 (7.8%) | 1193 (13.2%) | 7958 (11.9%) | 1204 (7.9%) |
| Diagnosis group (2+ Codes) | MDD-only | 787 (10.2%) | 1297 (5.2%) | 620 (6.1%) | 708 (10.1%) | 1005 (11.1%) | 5773 (8.6%) | 1177 (7.7%) |
|  | ANX-only | 373 (4.8%) | 2249 (9.1%) | 494 (4.9%) | 236 (3.4%) | 364 (4%) | 4170 (6.2%) | 643 (4.2%) |
|  | MDD+ANX | 684 (8.9%) | 3295 (13.3%) | 426 (4.2%) | 338 (4.8%) | 817 (9%) | 4947 (7.4%) | 728 (4.8%) |
| Current Age (yrs) | Mean (SD) | 76.5 (13.6) | 62.89 (16.31) | 60.4 (19.5) | 60.2 (15.5) | 61.2 (16.6) | 53 (22.6) | 40 (21.3) |
| Female | N (%) | 3811 (49.4%) | 13228 (53%) | 5148 (50.7%) | 4302 (61.1%) | 5511 (61%) | 39569 (59.1) | 9,032 (59.1) |
| EHR length (yrs) | Mean (SD) | 24.5 (14.3) | 12.95 (7.17) | 9.8 (2.1) | 10.2 (2.3) | 10.2 (2.8) | 10.7 (6.97) | 10.3 (7.3) |
| Total ICD codes | Mean (SD) | 596.7 (767.1) | 611.1 (741.4) | 126.2 (222.8) | 243.7 (344.9) | 238.7 (350.5) | 278.9 (390.8) | 255.5 (419.7) |

**eTable 4. PRS prediction of depression and anxiety separately.** Results are show for depression (MDD) and anxiety (ANX) defined from EHR with either 1+ codes required or 2+ codes required. R^2^ = percent variation explained on the liability scale.

|  |  |  |  | **1+ ICD codes** | | | **2+ ICD codes** | | |
| --- | --- | --- | --- | --- | --- | --- | --- | --- | --- |
|  | **Site** | **Ancestry** | **N.cont** | **N.case** | **R^2^** | **P-value** | **N.case** | **R^2^** | **P-value** |
| MDD | MayoGC | EUR | 5772 | 1947 | 0.9% | 3E-10 | 1644 | 1.0% | 2E-10 |
|  | MGB | EUR | 18077 | 6747 | 1.2% | 2E-40 | 5250 | 1.3% | 3E-37 |
|  | BioMe | EUR | 8614 | 1532 | 0.5% | 8E-06 | 1046 | 0.5% | 9E-05 |
|  | BioVU | EUR | 51644 | 15259 | 0.8% | 6E-65 | 10720 | 0.8% | 1E-50 |
|  | BioMe | AFR | 5463 | 1575 | 0.2% | 0.006 | 1046 | 0.1% | 0.070 |
|  | BioVU | AFR | 12607 | 2676 | 0.1% | 0.004 | 1905 | 0.2% | 0.0007 |
|  | BioMe | AMR | 6497 | 2537 | 0.4% | 8E-06 | 1822 | 0.6% | 4E-07 |
| ANX | MayoGC | EUR | 6210 | 1509 | 0.4% | 9E-05 | 1117 | 0.5% | 8E-05 |
|  | MGB | EUR | 16130 | 8694 | 0.4% | 5E-18 | 6108 | 0.7% | 3E-21 |
|  | BioMe | EUR | 8657 | 1489 | 0.1% | 0.050 | 920 | 0.1% | 0.087 |
|  | BioVU | EUR | 53373 | 13530 | 0.3% | 2E-24 | 9117 | 0.4% | 4E-20 |
|  | BioMe | AFR | 6086 | 952 | 0.0% | 0.46 | 574 | 0.0% | 0.44 |
|  | BioVU | AFR | 13206 | 2077 | 0.2% | 0.001 | 1371 | 0.2% | 0.013 |
|  | BioMe | AMR | 7248 | 1786 | 0.1% | 0.047 | 1181 | 0.1% | 0.13 |

**eTable 5. Joint PRS prediction of depression and anxiety in the same model.** Results are show for PRSs for depression (MDD) and anxiety (ANX) predicting anxiety and depression comorbid group compared to controls defined from EHR with 1+ codes required.

|  |  |  | MDD-PRS | | ANX-PRS | |
| --- | --- | --- | --- | --- | --- | --- |
| Site-Ancestry (N control) | Group | N case | OR (95% CI) | p-value | OR (95% CI) | p-value |
| Meta-EUR (N = 73296) | Anxiety-only | 10811 | 1.06 (1.04 ,1.09) | 2E-07 | 1.05 (1.03 ,1.08) | 7E-06 |
|  | Depression-only | 11074 | 1.12 (1.10 ,1.15) | 1E-23 | 1.05 (1.02 ,1.07) | 5E-05 |
|  | Comorbid | 14411 | 1.20 (1.18 ,1.23) | 1E-70 | 1.07 (1.05 ,1.09) | 2E-10 |
| Meta-AFR (N = 16792) | Anxiety-only | 1278 | 1.01 (0.94 ,1.07) | 0.83 | 1.03 (0.97 ,1.10) | 0.35 |
|  | Depression-only | 2500 | 1.07 (1.02 ,1.12) | 0.0092 | 0.98 (0.93 ,1.03) | 0.35 |
|  | Comorbid | 1751 | 1.07 (1.01 ,1.13) | 0.021 | 1.06 (1.00 ,1.12) | 0.053 |
| Bio*Me*-AMR (N = 5904) | Anxiety-only | 593 | 0.97 (0.89 ,1.07) | 0.54 | 1.08 (0.98 ,1.18) | 0.12 |
|  | Depression-only | 1344 | 1.07 (1.00 ,1.14) | 0.039 | 1.03 (0.96 ,1.10) | 0.39 |
|  | Comorbid | 1193 | 1.15 (1.08 ,1.24) | 4E-05 | 1.03 (0.96 ,1.10) | 0.44 |
| Mayo-EUR (N = 5216) | Anxiety-only | 556 | 1.04 (0.94 ,1.14) | 0.44 | 1.06 (0.96 ,1.17) | 0.24 |
|  | Depression-only | 994 | 1.18 (1.09 ,1.27) | 2E-05 | 1.03 (0.96 ,1.11) | 0.41 |
|  | Comorbid | 953 | 1.14 (1.06 ,1.23) | 8E-04 | 1.12 (1.04 ,1.20) | 0.0043 |
| MGB-EUR (N = 14208) | Anxiety-only | 3869 | 1.07 (1.03 ,1.12) | 3E-04 | 1.06 (1.02 ,1.10) | 0.0055 |
|  | Depression-only | 1922 | 1.17 (1.11 ,1.23) | 5E-09 | 1.05 (0.99 ,1.10) | 0.086 |
|  | Comorbid | 4825 | 1.22 (1.18 ,1.26) | < 1E-13 | 1.09 (1.05 ,1.13) | 1E-06 |
| Bio*Me*-EUR (N = 7800) | Anxiety-only | 814 | 1.12 (1.03 ,1.21) | 0.0047 | 1.02 (0.94 ,1.10) | 0.64 |
|  | Depression-only | 857 | 1.08 (1.00 ,1.16) | 0.049 | 1.06 (0.99 ,1.15) | 0.12 |
|  | Comorbid | 675 | 1.21 (1.11 ,1.31) | 1E-05 | 1.03 (0.94 ,1.12) | 0.52 |
| BioVU-EUR (N = 46072) | Anxiety-only | 5572 | 1.05 (1.01 ,1.08) | 0.0042 | 1.06 (1.02 ,1.09) | 6E-04 |
|  | Depression-only | 7301 | 1.11 (1.08 ,1.14) | 2E-12 | 1.05 (1.02 ,1.08) | 0.0011 |
|  | Comorbid | 7958 | 1.20 (1.17 ,1.24) | < 1E-13 | 1.05 (1.02 ,1.08) | 2E-04 |
| Bio*Me*-AFR (N = 5058) | Anxiety-only | 405 | 1.00 (0.89 ,1.11) | 0.942 | 1.01 (0.90 ,1.13) | 0.86 |
|  | Depression-only | 1028 | 1.08 (1.00 ,1.16) | 0.052 | 0.95 (0.89 ,1.03) | 0.23 |
|  | Comorbid | 547 | 1.13 (1.03 ,1.25) | 0.011 | 0.99 (0.90 ,1.09) | 0.83 |
| BioVU-AFR (N = 11734) | Anxiety-only | 873 | 1.01 (0.94 ,1.10) | 0.753 | 1.04 (0.96 ,1.13) | 0.308 |
|  | Depression-only | 1472 | 1.06 (0.99 ,1.13) | 0.077 | 0.99 (0.93 ,1.06) | 0.812 |
|  | Comorbid | 1204 | 1.04 (0.97 ,1.11) | 0.304 | 1.09 (1.02 ,1.17) | 0.012 |

**eTable 6. Joint PRS prediction of depression and anxiety in the same model.** Results are show for PRSs for depression (MDD) and anxiety (ANX) predicting anxiety and depression comorbid group compared to controls defined from EHR with 2+ codes required.

|  |  |  | MDD-PRS | | ANX-PRS | |
| --- | --- | --- | --- | --- | --- | --- |
| Site-Ancestry (N control) | Group | N case | OR (95% CI) | p-value | OR (95% CI) | p-value |
| Meta-EUR (N = 73296) | Anxiety-only | 8575 | 1.09 (1.06 ,1.12) | 3E-10 | 1.07 (1.04 ,1.09) | 5E-06 |
|  | Depression-only | 8477 | 1.13 (1.10 ,1.16) | 2E-21 | 1.06 (1.03 ,1.09) | 1E-05 |
|  | Comorbid | 9352 | 1.22 (1.19 ,1.26) | 1E-58 | 1.08 (1.05 ,1.10) | 4E-09 |
| Meta-AFR (N = 16792) | Anxiety-only | 879 | 1.00 (0.93 ,1.08) | 0.98 | 1.03 (0.96 ,1.12) | 0.38 |
|  | Depression-only | 1885 | 1.05 (0.99 ,1.10) | 0.11 | 1.01 (0.96 ,1.07) | 0.62 |
|  | Comorbid | 1066 | 1.12 (1.04 ,1.20) | 0.0016 | 1.04 (0.97 ,1.12) | 0.25 |
| Bio*Me*-AMR (N = 5904) | Anxiety-only | 364 | 1.02 (0.91 ,1.15) | 0.68 | 1.06 (0.94 ,1.19) | 0.36 |
|  | Depression-only | 1005 | 1.11 (1.03 ,1.19) | 0.0059 | 1.04 (0.97 ,1.12) | 0.28 |
|  | Comorbid | 817 | 1.19 (1.10 ,1.29) | 1E-05 | 1.01 (0.94 ,1.10) | 0.73 |
| Mayo-EUR (N = 5216) | Anxiety-only | 373 | 0.99 (0.88 ,1.12) | 0.91 | 1.10 (0.98 ,1.23) | 0.12 |
|  | Depression-only | 787 | 1.18 (1.09 ,1.28) | 8E-05 | 1.05 (0.97 ,1.14) | 0.245 |
|  | Comorbid | 684 | 1.18 (1.08 ,1.29) | 2E-04 | 1.13 (1.04 ,1.23) | 0.006 |
| MGB-EUR (N = 14208) | Anxiety-only | 2249 | 1.08 (1.03 ,1.14) | 0.0016 | 1.09 (1.03 ,1.14) | 0.0010 |
|  | Depression-only | 1297 | 1.17 (1.10 ,1.24) | 9E-07 | 1.05 (0.98 ,1.11) | 0.15 |
|  | Comorbid | 3295 | 1.25 (1.19 ,1.30) | < 1E-13 | 1.10 (1.06 ,1.15) | 5E-06 |
| Bio*Me*-EUR (N = 7800) | Anxiety-only | 494 | 1.19 (1.08 ,1.32) | 4E-04 | 1.02 (0.93 ,1.13) | 0.64 |
|  | Depression-only | 620 | 1.07 (0.98 ,1.17) | 0.14 | 1.06 (0.97 ,1.16) | 0.2 |
|  | Comorbid | 426 | 1.27 (1.14 ,1.41) | 1E-05 | 1.00 (0.90 ,1.11) | 0.95 |
| BioVU-EUR (N = 46072) | Anxiety-only | 4170 | 1.09 (1.05 ,1.13) | 2E-06 | 1.06 (1.02 ,1.10) | 0.0027 |
|  | Depression-only | 5773 | 1.12 (1.09 ,1.16) | 4E-13 | 1.06 (1.03 ,1.10) | 1E-04 |
|  | Comorbid | 4947 | 1.21 (1.17 ,1.25) | < 1E-13 | 1.06 (1.03 ,1.10) | 7E-04 |
| Bio*Me*-AFR (N = 5058) | Anxiety-only | 236 | 1.00 (0.86 ,1.15) | 0.964 | 1.01 (0.87 ,1.16) | 0.94 |
|  | Depression-only | 708 | 1.02 (0.94 ,1.11) | 0.641 | 1.02 (0.94 ,1.12) | 0.61 |
|  | Comorbid | 338 | 1.15 (1.02 ,1.30) | 0.027 | 1.01 (0.89 ,1.14) | 0.92 |
| BioVU-AFR (N = 11734) | Anxiety-only | 643 | 1.00 (0.91 ,1.10) | 0.953 | 1.05 (0.96 ,1.15) | 0.33 |
|  | Depression-only | 1177 | 1.06 (0.99 ,1.14) | 0.096 | 1.01 (0.94 ,1.08) | 0.82 |
|  | Comorbid | 728 | 1.11 (1.01 ,1.21) | 0.022 | 1.06 (0.97 ,1.15) | 0.19 |

### **Supplementary References**

1.	Bielinski SJ, Chai HS, Pathak J, et al. Mayo Genome Consortia: a genotype-phenotype resource for genome-wide association studies with an application to the analysis of circulating bilirubin levels. *Mayo Clin Proc*. 2011;86(7):606-614.

2.	Das S, Forer L, Schönherr S, et al. Next-generation genotype imputation service and methods. *Nat Genet*. 2016;48(10):1284-1287.

3.	Belbin GM, Cullina S, Wenric S, et al. Toward a fine-scale population health monitoring system. *Cell*. 2021;184(8):2068-2083.e11.

4.	Howie BN, Donnelly P, Marchini J. A flexible and accurate genotype imputation method for the next generation of genome-wide association studies. *PLoS Genet*. 2009;5(6):e1000529.

5.	Dennis J, Sealock J, Levinson RT, et al. Genetic risk for major depressive disorder and loneliness in sex-specific associations with coronary artery disease. *Mol Psychiatry*. 2021;26(8):4254-4264.

6.	Delaneau O, The 1000 Genomes Project Consortium, Marchini J. Integrating sequence and array data to create an improved 1000 Genomes Project haplotype reference panel. *Nature Communications*. 2014;5(1). doi:[10.1038/ncomms4934](http://dx.doi.org/10.1038/ncomms4934)
